# HIV post-exposure prophylaxis adherence due to needle stick and sharp injuries

**DOI:** 10.1101/2023.04.25.23289085

**Authors:** Moses Odhiambo Osoo, George Ochieng’ Otieno, Peter Halestrap

**Affiliations:** Department of Research, AIC Kijabe Hospital, Kijabe, Kenya; Department of Internal Medicine, AIC Kijabe Hospital, Kijabe, Kenya; Department of Family Medicine, AIC Kijabe Hospital, Kijabe, Kenya

**Keywords:** HIV Post-Exposure Prophylaxis, Blood-Borne Pathogens, Needlestick Injuries, Health Personnel, Occupational Health

## Abstract

**Background:** Healthcare workers frequently experience needle sticks and other sharp injuries, which are among the most prevalent types of occupational accident in hospital settings. Studies indicate that approximately 50% of these devices are contaminated with blood, exposing users to potential infections from blood-borne pathogens such as HIV. Despite the efficacy of post-exposure prophylaxis in lowering the risk of HIV transmission, ensuring consistent adherence to this treatment regimen remains a widely acknowledged challenge.

**Objective:** This study aimed to assess healthcare workers’ adherence to HIV post-exposure prophylaxis following needlestick and sharp injuries at Kijabe Hospital, Kenya, between February 2017 and June 2022.

**Method:** A retrospective chart review was conducted on healthcare workers who were started on HIV PEP following a reported occupational exposure due to needle sticks and sharp injuries between February 2017 and June 2022.

**Result:** In total, 136 healthcare workers were exposed to needle sticks and sharp injuries. The majority of exposures (82.3%) were at a high risk of HIV transmission. The overall adherence rate to the 28-day HIV-PEP course was 26%. Healthcare workers were more likely to complete HIV-PEP when the source’s HIV status was positive than when it was negative [ 42.4% vs. 11.8%; p = 0.001]. The drug regimen (TDF/3TC/DTG) was significantly associated with default treatment (aOR, 5.0 (95% CI 2.0 – 11.0) (p= 0.001) compared with patients using the TDF/3TC/ATVr regimen.

**Conclusion:** Our study found that 74% of healthcare workers did not complete the full 28-day course of HIV post-exposure prophylaxis (PEP) as recommended, indicating a significant issue with adherence. This highlights the need for strategies to improve compliance and ensure occupational health. Additionally, as there is limited research on HIV PEP completion rates among healthcare workers in Kenya, these findings will contribute valuable knowledge in this area.

## Study Background

Needle stick and sharp injuries (NSSIs) among healthcare workers (HCWs) are among the most common occupational accidents in hospitals^1^. Approximately half of the instruments responsible for these injuries are contaminated with blood, putting users at risk of blood-borne pathogens such as hepatitis B, hepatitis C, and HIV^2–4^. HIV remains a major global public health concern, having claimed more than 35 million lives to date. Africa is the most affected region worldwide, with 25.7 million HIV-positive people. Recently, the region has accounted for nearly two-thirds of all new HIV infections, and these factors make healthcare workers in this region at a particularly increased risk for HIV transmission following a needlestick injury ^5^; this is exacerbated by the fact that the prevalence of NSSIs among HCWs is higher in developing countries than in developed countries^6^. It is estimated that 4.4% of HIV infections and 39% of HBV and HCV infections are attributed to occupational injury^7^. The risk of HIV transmission is estimated to be 0.3% after a needle-stick injury and 0.09% after mucous membrane exposure^8^. Deeper injuries, as well as the visibility of blood on sharp devices, are associated with an increased risk of HIV transmission ^9^.

In Kenya, guidelines recommend that healthcare workers (HCWs) should start post-exposure prophylaxis (PEP) following high-risk exposure. This includes exposure to a large volume of blood or potentially infectious fluids or exposure to blood or body fluids contaminated with blood from a patient with a high viral load. Additionally, a deep and extensive injury from a contaminated sharp instrument or exposure to blood from an HIV drug-resistant patient also necessitates PEP, according to the guidelines^10^. The primary risk of disease transmission is determined by the type of exposure and specific materials involved. This type of exposure can include contact with mucous membranes (such as through sexual activity or contact with splashes to the eye, nose, or mouth), non-intact skin, percutaneous injury, or parenteral exposure. Meanwhile, materials that pose a risk for transmission include blood, blood-stained body fluids, breast milk, semen, vaginal secretions, synovial fluid, pleural fluid, pericardial fluid, amniotic fluid, cerebrospinal fluid, and HIV cultures in laboratory settings.

Adherence to and completion of PEP treatment strategies are important for maximizing HCWs risk reduction; however, adherence has been shown to be poor in many contexts^11–15^. HIV PEP utilization is lower in sub-Saharan Africa (SSA) than in other parts of the globe^16,17^ despite the generally higher prevalence of HIV in this setting. This study aimed to assess adherence to HIV PEP after needlestick and sharp injuries among healthcare workers at Kijabe Hospital, Kenya, between February 2017 and June 2022.

## Materials and Methods

### Study Design

A retrospective chart review was conducted on HCWs who started taking HIV PEP following a reported occupational exposure due to NSSIs between February 2017 and June 2022 to assess completion of the 28-day course.

### Setting

Kenyan peri-urban, tertiary care center referral hospital in central Kenya with a 360-bed. At the time of the study, the hospital had 869 HCWs and saw approximately 600 patients per day in an outpatient setting, in addition to performing over 9000 surgeries per year.

### Study Population

A census of all reported occupational exposure due to NSSIs among Kijabe Hospital HCWs who were started on HIV PEP between February 2017 and June 2022.

### Data Collection

Data were extracted from the Ministry of Health HIV-PEP registry without patient identifiers. Data quality checks were performed to ensure consistency and completeness of the data. The variables included HCWs characteristics, risk of infection, location of exposure, treatment regimen, and treatment completion. Variables of interest: outcome (adherence), exposure (Started on HIV PEP), predictors (risk of infection, HIV PEP regimen), and potential confounders (adverse drug reactions). Potential sources of bias (selection bias was unavoidable; only data of HCWs who reported exposure and started HIV PEP were considered for analysis). Data were extracted in June, 2022.

### Data Analysis

Healthcare workers characteristics were assessed using descriptive statistics. Chi-square or Fisher’s exact tests were used for bivariate analyses. Adherence was calculated by dividing the number of participants who completed an entire 28-day course of HIV PEP by the number of participants who started on HIV PEP following occupational exposure due to NSSIs. Logistic regression was used to assess the predictors of HIV-PEP adherence at a 95% confidence interval. Missing values were removed by list-wise deletion from the variable of interest for the logistic regression model. Data were analyzed using STATA version 18.

### Ethical Consideration

Ethical waiver was sought from an independent ethics committee [**Tenwek Hospital ISERC (Institutional Scientific and Ethical Review Committee) approval number – 2023-0015**]

No informed consent was required since this was a retrospective chart review

### Data confidentiality

Clinicians in the Comprehensive Care Clinic (CCC) involved in routine HIV care retrieved anonymized data for review and data analysis. The authors had no access to information that could identify individual participants during or after the data collection. The data collected were pseudonymized in compliance with the hospital’s data protection policy.

## Data Availability

The datasets generated during and/or analyzed during the current study are available in the Dryad^18^ at https://doi.org/10.5061/dryad.0cfxpnw78

### Disclaimer

The views and opinions expressed in this article are those of the authors and do not necessarily reflect the official policies or positions of affiliated institutions of the authors.

## Results

### Demographic characteristics of healthcare workers

A total of 136 exposures to NSSIs have been reported. The majority (85.7%) of the HCWs were between the ages of 18 and 35 years, and more than half (54.4%) were male. Approximately 73% of these HCWs were nurses, doctors, and students. Thirty-nine percent of the exposures were reported from the theatre, followed by the medical-surgical ward at 29.6%. As shown in Table I

**Table I:** Healthcare worker characteristics started on HIV PEP After NSSIs.

| Variable | Completed<br>(n=35,25.7%) | Defaulted<br>(n=100,73.5%) | Declined<br>(n=1,0.7%) | Total (N=136) | P-value |
| --- | --- | --- | --- | --- | --- |
| **Age in yrs, median (iqi)** | 28.00 (25.00; 32.25) | 26.50 (23.00; 32.00) | 37.00 (37.00; 37.00) | 28.00 (23.50; 32.00) | 0.20 |
| **Age category(years)\*, n (%)** |  |  |  |  | 0.14 |
| 18 - 35 years | 29 (25.4) | 85 (74.6) | 0 (0.0) | 114 (85.7) |  |
| 36-59 years | 5 (27.8) | 12 (66.7) | 1 (5.6) | 18 (13.5) |  |
| 60 years and above | 0 (0.0) | 1 (100.0) | 0 (0.0) | 1 (0.8) |  |
| **Sex, n (%)** |  |  |  |  | 0.52 |
| Male | 20 (27.0) | 54 (73.0) | 0 (0.0) | 74 (54.4) |  |
| Female | 15 (24.2) | 46 (74.2) | 1 (1.6) | 62 (45.6) |  |
| **Year of exposure, n (%)** |  |  |  |  | 0.25 |
| 2017 | 6 (31.6) | 13 (68.4) | 0 (0.0) | 19 (14.0) |  |
| 2018 | 10 (28.6) | 24 (68.6) | 1 (2.9) | 35 (25.7) |  |
| 2019 | 9 (39.1) | 14 (60.9) | 0 (0.0) | 23 (16.9) |  |
| 2020 | 4 (14.8) | 23 (85.2) | 0 (0.0) | 27 (19.9) |  |
| 2021 | 5 (38.5) | 8 (61.5) | 0 (0.0) | 13 (9.5) |  |
| 2022 | 1 (5.3) | 18 (94.7) | 0 (0.0) | 19 (14.0) |  |
| **Location during exposure<sup>#</sup>, n (%)** |  |  |  |  | 0.97 |
| Medical-Surgical | 13 (32.5) | 27 (67.5) | 0 (0.0) | 40 (29.6) |  |
| Theatre | 12 (22.6) | 40 (75.5) | 1 (1.9) | 53 (39.3) |  |
| Maternity | 2 (16.7) | 10 (83.3) | 0 (0.0) | 12 (8.8) |  |
| OPD/A&E | 4 (30.8) | 9 (69.2) | 0 (0.0) | 13 (9.6) |  |
| Laboratory | 1 (20.0) | 4 (80.0) | 0 (0.0) | 5 (3.8) |  |
| Laundry | 0 (0.0) | 3 (100.0) | 0 (0.0) | 3 (2.2) |  |
| others | 2 (22.2) | 7 (77.8) | 0 (0.0) | 9 (6.7) |  |
| **Duration before STAT dose\*\*, n (%)** |  |  |  |  |  |
| <72hrs | 31 (26.5) | 85 (72.6) | 1 (0.9) | 117 (100) | . |

\*A denominator of 133 was used to calculate total percentages, missing values (3)

### Adherence to HIV PEP among exposed healthcare workers due to NSSIs

The overall adherence rate to the 28-day HIV-PEP course was 26%. Eighty-two percent of the exposures were high-risk. HCWs were more likely to complete HIV PEP when the HIV status of the source was positive compared to when the HIV status of the source was negative **[ 42.4% vs 11.8%; p = 0.001**]. The HIV status of the source patient/specimen was reported in 49% of cases. Variables with a p value <.09 in univariate analysis were included in multivariate logistic regression analysis (Risk of HIV infection and HIV PEP regimen). Summary on table II

**Table II:** Assessing factors associated to HIV PEP adherence among HCWs following needle stick and sharp injuries; 2017-2022.

| Adherence | Adhered<br>(n=35,25.7%) | Defaulted<br>(n=101,74.3%) | Total (N=136) | P-value |
| --- | --- | --- | --- | --- |
| **Age in yrs, median (iqi)** | 28.00 (25.00;<br>32.25) | 27.00 (23.00; 32.00) | 28.00 (23.50;<br>32.00) | 0.26 |
| **Sex, n (%)** |  |  |  | 0.71 |
| Male | 20 (27.0) | 54 (73.0) | 74 (54.4) |  |
| Female | 15 (24.2) | 47 (75.8) | 62 (45.6) |  |
| **HIV status <sup>□</sup>, n (%)** |  |  |  | 0.00 |
| Negative | 4 (11.8) | 30 (88.2) | 34 (50.7) |  |
| Positive | 14 (42.4) | 19 (57.6) | 33 (49.3) |  |
| **Risk of HIV infection <sup>□</sup>, n (%)** |  |  |  | 0.07 |
| Low | 9 (39.1) | 14 (60.9) | 23 (17.7) |  |
| High | 23 (21.5) | 84 (78.5) | 107 (82.3) |  |
| **HIV Regimen<sup>¶</sup>, n (%)** |  |  |  | 0.00 |
| TDF/3TC/ATVr | 23 (44.2) | 29 (55.8) | 52 (39.4) |  |
| TDF/3TC/DTG | 12 (15.2) | 67 (84.8) | 79 (59.8) |  |
| AZT/3TC/LOPr | 0 (0.0) | 1 (100.0) | 1 (0.8) |  |

– A denominator of 67 (Known HIV status of source/specimen) was used to calculate total percentages
– A denominator of 130 was used to calculate total percentages, missing values (6)
<sup>¶</sup>A denominator of 132 was used to calculate total percentages, missing values (3)

### Factors associated with HIV PEP adherence among 128 HCWs following needle stick and sharp injuries; 2017-2022

Multivariate logistic regression was used to assess the effect of the HIV PEP regimen and risk of infection on adherence to the 28-day course. HIV PEP regimen was significantly associated with adherence in bivariate and multivariate analyses: cOR; 5.0 (95% CI 2.1 – 12.0) (p=0.001) and aOR; 4.7 (95% CI 2.0 – 11.0) (p= 0.001). The drug regimen (TDF/3TC/DTG) was significantly associated with non-adherence, with patients using the TDF/3TC/ATVr regimen having a higher completion rate. Further details are provided in Table III.

**Table III:** Unadjusted and adjusted Odds ratios from logistic regression analysis.

| **Risk factors** | **Unadjusted Odds Ratio**<br>**[95% CI]** | **p-value** | **Unadjusted Odds**<br>**Ratio [95% CI]** | **p-value** |
| --- | --- | --- | --- | --- |
| **Risk of Hiv Infection** |  |  |  |  |
| High | 2.3[0.88;6.0] | 0.089 | 1.7[0.59;4.6] | 0.337 |
| Low | ref |  |  |  |
| **HIV PEP Regimen** |  |  |  |  |
| TDF/3TC/ATVr | ref |  |  |  |
| TDF/3TC/DTG | 5.0[2.1;12.0] | 0.001 | 4.7[2.0;11.0] | 0.001 |
| AZT/3TC/LPVr | ref |  |  |  |

### Discussion

Adherence to 28-day HIV post-exposure prophylaxis (PEP) among healthcare workers (HCWs) following occupational exposure due to needlestick and sharps injuries (NSSIs) was found to be only 26%. This low adherence rate is concerning, especially considering that over 80% of the exposures among these HCWs were categorized as high-risk. This adherence rate aligns with a previous study by Kimaro et al., which reported an adherence rate of 23.1%^19^, but it is significantly lower than that reported in other studies conducted in Botswana, Ghana, and South Africa. In those studies, adherence to HIV PEP was 71%, 69%, and 55.6%, respectively^17,20–22^. This significant variation in adherence rates across different regions requires further investigation to understand the underlying factors that contribute to this disparity.

According to the guidelines of the Kenya Ministry of Health^10^, it is recommended that HIV post-exposure prophylaxis (PEP) be completed regardless of the HIV status of the source material because of the high HIV prevalence in these settings. This study revealed that healthcare workers were more likely to follow through with HIV PEP when the source’s HIV status was positive as opposed to when it was negative. This difference in completion rates could be linked to a heightened concern about seroconversion when the source is HIV-positive, indicating that these healthcare workers possessed sufficient knowledge about HIV PEP. These findings align with those of a study conducted by Owolabi et al., which reported poor knowledge and practice of PEP among healthcare providers. The study found that many healthcare providers who were exposed to HIV-positive sources did not initiate PEP^23^.

The study also found that the HIV PEP drug regimen was significantly associated with adherence rates, and intolerance to adverse events has been cited as the sole reason for discontinuing HIV PEP^24,25^, which underlines the necessity for comprehensive and effective counseling, education, proactive follow-up (which could involve mobile or phone contact), and management of adverse events. Providing education on the importance of completing the PEP schedule, particularly for healthcare workers on a 28-day schedule, can significantly enhance adherence, which is crucial for minimizing the risk of HIV seroconversion.

The HIV PEP treatment protocol places strong emphasis on treatment completion. It encompasses a comprehensive approach that starts with prompt recognition, providing first aid, and reporting events, in addition to initiating antiretroviral therapy (ARVs). We encountered challenges in tracking healthcare workers (HCWs) who were exposed to HIV, but did not seek PEP treatment. As a result, we may not have accounted for other factors that influence adherence to PEP protocols in general.

## Conclusion

In our study, we found that only 26% of healthcare workers (HCWs) completed HIV post-exposure prophylaxis (PEP) after experiencing needle-stick and sharp object injuries (NSSIs). We also found a significant association between adherence and HIV PEP regimen. Our research highlights the need for further investigation into the reasons for HCWs defaulting on PEP with the goal of developing targeted strategies to improve adherence and enhance occupational health. The findings of our study are not only relevant to healthcare workers in our specific facility but also have implications for healthcare workers in other settings across Kenya and sub-Saharan Africa.

## Acknowledgements

Mary B. Adam –Manuscript review, Wilbon Korir-Data collection

